# Early detection of patients with narcotic use disorder using a modified MEDD score based on the analysis of real-world prescription patterns

**DOI:** 10.1101/2022.04.12.22273679

**Authors:** Yi-Jun Kim, Kye Hwa Lee, Hye-Ryun Kang, Yoon Sook Cho, Dong Yoon Kang, Ju Han Kim, Yon Su Kim

## Abstract

**Background:** Addiction to prescription narcotics is a global issue, and detection of individuals with a narcotic use disorder (NUD) at an early stage can help prevent narcotics misuse and abuse. We developed a novel index to detect early NUD based on a real-world prescription pattern analysis in a large hospital.

**Methods:** We analyzed narcotic prescriptions of 221 887 patients prescribed by 8737 doctors from July 2000 to June 2018. For the early detection of patients who could potentially progress to developing NUD after a long history of narcotic prescription, a weighted morphine equivalent daily dose (wt-MEDD) score was developed based on the number of prescription dates on which the actual MEDD was higher than the intended MEDD. Performance of the wt-MEDD scoring system in detecting patients diagnosed with NUD by doctors was compared with that of other NUD high risk indexes such as the MEDD scoring system, number of days on prescribed narcotics, the frequency/duration of prescription, narcotics prescription across multiple doctors, and the number of early refills of narcotics.

**Results:** A wt-MEDD score cut-off value of 10.5 could detect all outliers, as well as patients diagnosed with NUD with 100% sensitivity and 99.6% specificity. The wt-MEDD score showed the highest sensitivity and specificity in identifying NUD among all indexes. Further, combining the wt-MEDD score with other NUD high risk indexes improved the prediction performance.

**Conclusion:** We developed a novel index to distinguish patients with vulnerable use patterns of narcotics. The wt-MEDD score showed excellent performance in detecting early NUD.

## INTRODUCTION

According to the World Drug Report, 2020 ^1^, the number of deaths attributed to opioid overdose increased 2.5-fold from 18,515 deaths in 2007 to approximately 47,000 deaths in 2018. A prominent feature of the current narcotic overdose crisis is the increased prevalence of addiction to prescription narcotics ^2^. Among patients who are prescribed narcotics owing to chronic pain, 21–29% of them misuse it and 8–12% progress to narcotics addiction ^3^. Public awareness of the seriousness of addiction to prescription narcotics has been rising in recent years ^4^.

Narcotic abuse can be defined in various ways; for example, MedlinePlus defines prescription narcotic abuse as “taking medicine in a way that is different from what the doctor prescribed” ^5^. Narcotics abuse is mostly caused by a desire to obtain more narcotics than those prescribed by doctors owing to a “strong desire or urge to use the substance” ^6^. To prevent prescription narcotic use disorder (NUD), the Centers for Disease Control and Prevention (CDC) of the United States has provided a guideline for prescribing narcotics ^7^. Most states are now required to register narcotic users through prescription drug monitoring programs (PDMP), which monitor narcotic prescriptions ^8^. According to the CDC guideline, prescribing a morphine equivalent daily dose (MEDD), or morphine milligram equivalent (MME) per day of ≥ 90 should be restricted as much as possible ^7^. Some studies have suggested that this system has reduced NUD prevalence and the associated side effects ^9,10^; however, a recent review has shown ambiguous results ^11^.

A limitation of the CDC guideline and PDMP is that these can only detect the risk of NUD, and not NUD itself. In addition, the cut-off values of the NUD high risk indexes are not absolute standards to identify patients vulnerable to NUD. For example, the restriction cut-off of MEDD shows wide variability across multiple countries: 90 MME/day in the USA and 200 MME/day in Canada ^12^. Many studies have attempted to develop tools to predict NUD; however, these tools are mainly focused on analyzing the high-risk factors of NUD and not on the actual detection of NUD ^13,14^. Thus, a method for detecting an absolute abnormal prescription pattern of NUD has to be developed.

Hence, in this study, we devised a method for screening patients with NUD based on the definition of prescription narcotic abuse itself via exploring large real-world clinical data. Multiple strategies adopted by a patient to receive more narcotics could result in the actual MEDD of a patient being higher than that intended by doctors because the MEDDs overlap. We developed a weighted (wt)-MEDD score based on the number of prescription dates on which the increase in the MEDD ratio [(actual MEDD)/(intended MEDD)] was above a certain level (for example, 1.5), indicating the presence of NUD based on the definition of prescription narcotics abuse. We evaluated the usefulness of this wt-MEDD score in detecting patients diagnosed with NUD by doctors at the early stage of the narcotic prescription path by comparing its performance with that of other narcotic prescription-related indexes.

## METHODS

### Study setting and population

This study assessed the total narcotics prescriptions from July 2000 to June 2018 in a single large hospital. Narcotics prescribed to patients with cancer and to inpatients were excluded from the analysis. The following 12 narcotics were selected for analysis: fentanyl, hydrocodone, hydromorphone, morphine, oxycodone, oxycodone/naloxone, tapentadol, alfentanil, meperidine, remifentanil, buprenorphine, and nalbuphine. Low-dose narcotics, such as codeine, and tramadol were excluded from the analysis.

A clinical data warehouse is a near-real-time database that consolidates data from various clinical sources. A web-based browser was used to extract the list of patients who met the inclusion criteria, and their corresponding electronic medical records were extracted. Information on the prescribed patients was downloaded from the clinical data warehouse into five tables: basic information, narcotic prescription, admission, surgery, and diagnosis record. This study was approved by the Institutional Review Board (IRB) of the Seoul National University Hospital (IRB No. 1806-182-955) and was performed in accordance with the ethical standards laid down in the 1964 Declaration of Helsinki.

### Design

This retrospective cohort study utilized real-world data to devise detection methods based on a specific definition of narcotic abuse. Most patterns of NUD involve taking higher doses than that intended by doctors. The hypothesis was that a patient at risk of developing NUD will try multiple strategies for higher MEDD, resulting in a difference between the MEDD that the doctors intended to prescribe to the patient and the overlapping MEDD that the patient achieves by utilizing these strategies. Therefore, the MEDD ratio is defined as follows (Fig. 1-a):

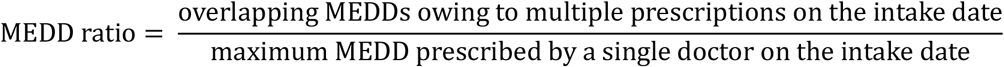

**Fig. 1.**
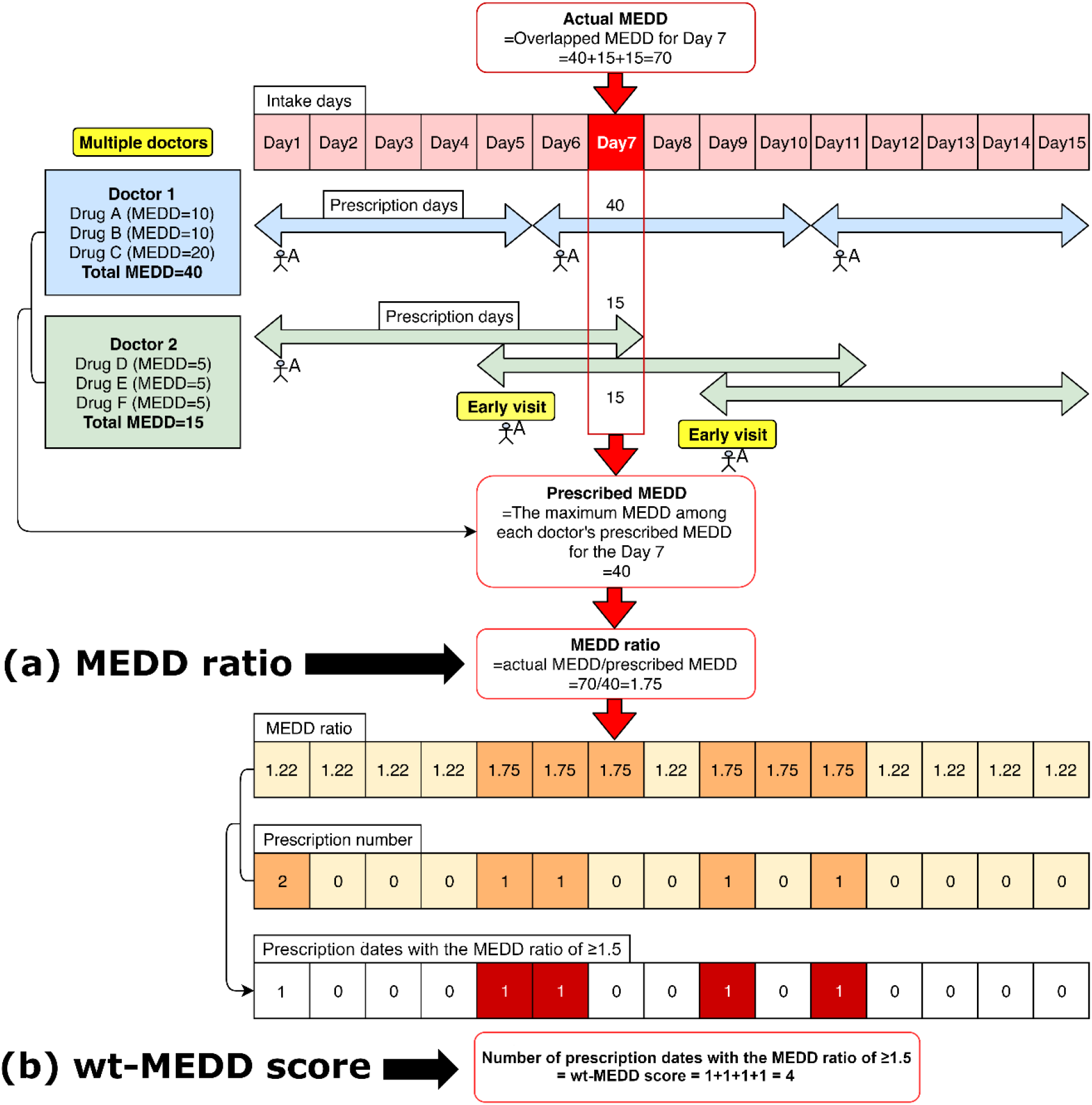
Schematic diagram of the calculation of weighted MEDD score. MEDD, morphine equivalent daily dose; wt-MEDD, weighted MEDD.

For example, doctor 1 prescribes 40 MEDD to patient A, considering that 40 MEDD is an appropriate amount for patient A, but patient A visits doctor 2 again to receive an additional narcotics prescription. Doctor 2 is unaware of the prescription of doctor 1 and prescribes 15 MEDD as the appropriate dose for patient A. However, patient A revisits doctor 2 earlier than the scheduled date for the next visit and receives the same prescription again, saying that he has lost his previously prescription. As a result, patient A receives a total of 40 + 15 + 15 MEDD of narcotics, which is 1.75 times higher than the intended 40, the highest MEDD prescribed by doctors to patient A. The MEDD ratio is defined as a ratio between the actual MEDD and the maximum MEDD of the intended prescription by doctors. In this case, the MEDD ratio is 1.75 (Fig. 1-a).

In addition, we defined the wt-MEDD score as follows (Fig. 1-b):

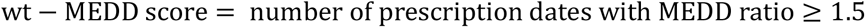

In Figure 1, patient A visits doctor 1 on days 1, 6, and 11, and doctor 2 on days 1, 5, and 9. Therefore, the total number of prescription dates for patient A is 5 (Days 1, 5, 6, 9, and 11). Among the five prescription dates, the number of dates with the MEDD ratio of ≥ 1.5 is 4 (Day 5, 6, 9, and 11). The wt-MEDD score is defined as the number of prescription dates with the MEDD ratio of ≥ 1.5. In this case, the wt-MEDD score is 4 (Fig. 1-b). If patient A continues the pattern of visiting multiple doctors for narcotics prescription and visiting doctors earlier than the scheduled date, the wt-MEDD score will increase continuously.

The reason for choosing a MEDD ratio of 1.5 (more than 1.0 but less than 2.0) was to allow acceptable minor differences between doctors’ intended MEDD prescription and actual MEDD (for example, in a situation of adjusting the appropriate dose at the beginning of the narcotics prescription) while sufficiently detecting abnormal prescriptions that need to be reviewed. MEDD ratios less than 1.5 might be too sensitive to detect meaningful inconsistencies between actual MEDD and intended MEDD. The cut-off MEDD ratio can be modified according to the preference of the institutions (less than 1.5 for higher sensitivity or more than 1.5 for higher specificity). The wt-MEDD score was used as a surrogate for NUD because repeated prescriptions with a high MEDD rate imply that the patient is repeatedly taking more narcotics than the doctor had intended to prescribe.

We explored whether the wt-MEDD score can be used clinically. First, to monitor abnormal prescription patterns of narcotics in a hospital, it is necessary to identify the doctors and patients involved in these practices and provide feedback to them. We created a list of doctors and patients related to abnormal prescription patterns using the wt-MEDD score. We generated a list of doctors who had outlier wt-MEDD scores. A list of patients with outlier wt-MEDD scores was also generated to monitor their prescription patterns. Such doctors and patients were defined as those who satisfied a two-tailed p-value of < 0.001 (Z score of ≥ 3.29 or ≤ -3.29). The list of doctors and patients with outlier wt-MEDD scores was extracted and the cut-off wt-MEDD score for both was obtained, to monitor abnormal narcotics prescription patterns.

Second, we examined whether the wt-MEDD score could be used to detect NUD patients at an earlier stage. We analyzed the optimal cut-off value of the wt-MEDD score to detect patients who were diagnosed with NUD by doctors, with the highest sensitivity and specificity. If the cut-off of the wt-MEDD score showed high sensitivity and specificity for a patient diagnosed with NUD, and if the time point at which this cut-off value was reached was earlier than the time of NUD diagnosis by doctors, this cut-off value can be used as an indicator to detect NUD at an early stage. The list of patients diagnosed with NUD by doctors was extracted from the clinical data warehouse using the 10^th^ revision of the International Statistical Classification of Disease and Related Health Problems (ICD-10) code and diagnostic terms in the doctors’ medical chart. The accuracy of the diagnosis of NUD was confirmed by checking whether the contents of the chart record satisfied the diagnostic criteria of NUD according to the Diagnostic and Statistical Manual of Mental Disorders, Fourth Edition, Text Revision (DSM-VI-tr) or DSM-V. Only those patients who were repeatedly prescribed narcotics in our hospital before being diagnosed with NUD were selected. Patients who were previously diagnosed with NUD from other hospitals with little or no history of narcotics prescription in our hospital were excluded. Two doctors reviewed the electronic medical record charts to confirm the accuracy of the diagnoses (Fig. 2). Prescriptions for these patients after the diagnosis of NUD were excluded from the analysis.

**Fig. 2.**
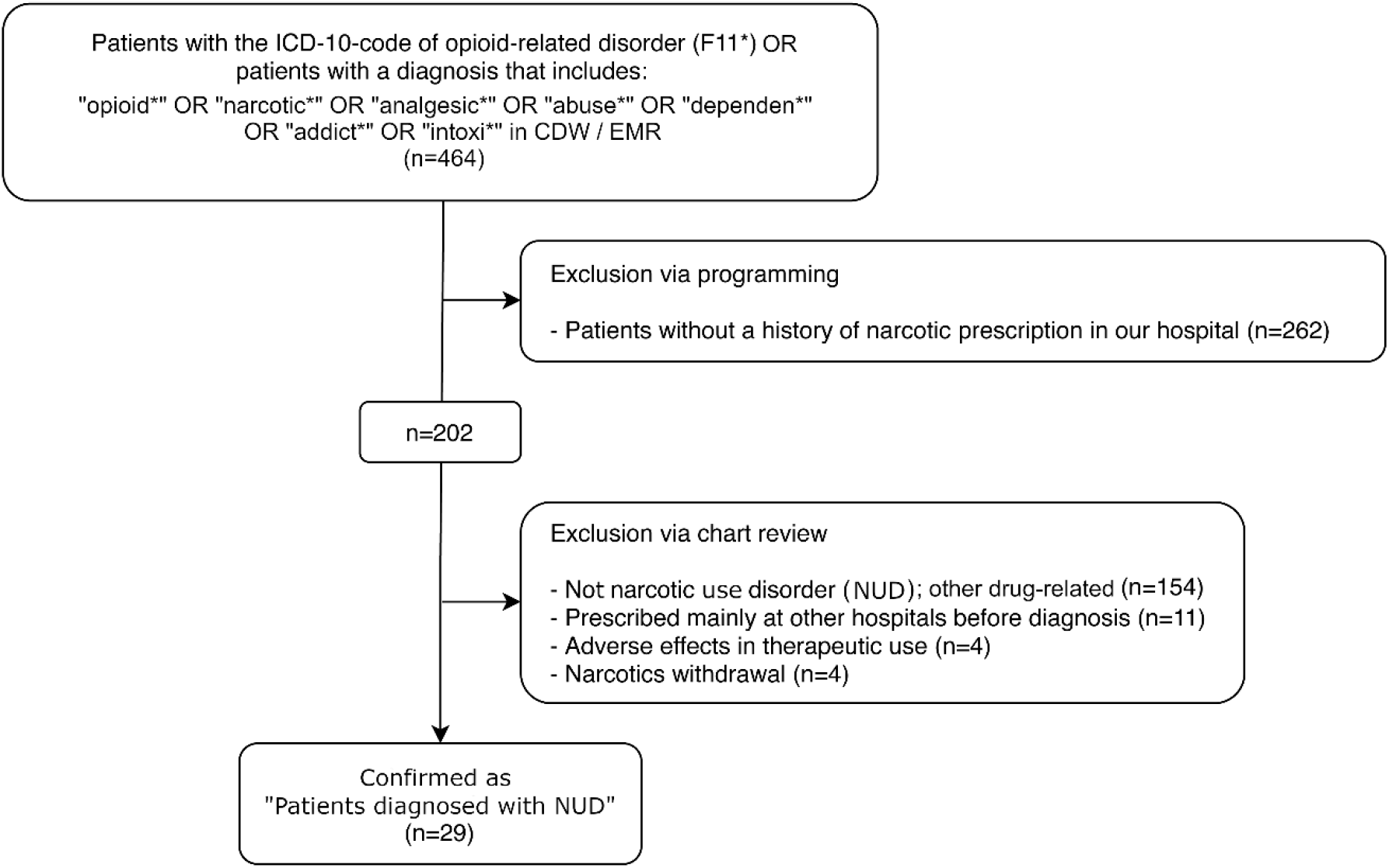
Flowchart of patient screening and enrollment. ICD, international classification of diseases; CDW, clinical data warehouse; EMR, electronic medical records.

We also analyzed the optimal cut-off values and the sensitivity and specificity of other NUD high risk indexes (such as the PDMP monitoring categories) and their combinations to confirm the usefulness of the wt-MEDD score. A McNemar test was performed between the wt-MEDD score and other indexes to determine whether the differences in the sensitivity and specificity were statistically significant.

We observed the time points of reaching the cut-off values of the wt-MEDD score and other NUD high risk indexes, and the time points of NUD diagnosis by a doctor in a patient case to investigate whether the cut-off value of wt-MEDD score could be used to identify NUD earlier. Paired t-tests were used to compare the mean time from the first prescription of narcotics to reaching the cut-off value of the wt-MEDD score and the NUD diagnosis by doctors.

### Data analysis

Considering the definition of narcotic abuse and the PDMP monitoring conditions, the wt-MEDD score and NUD high risk indexes (MEDD, prescription days, prescribing frequency and duration, number of prescribing doctors, and number of early receipt of narcotics before the scheduled visit) were selected for analysis.

We calculated the activity of each narcotic according to its mode of administration (tablet, patch, or injection). Information on the MME conversion factor was included in a table based on PDMP supplements ^15^. MEDD corresponds to the multiplication of the MME conversion factor and daily dose; when the MME conversion factor information for a certain drug was unavailable, it was estimated from the relevant literature ^16^.

Among the five types of downloaded tables, the narcotic prescription table contained drug information such as prescribed drug name, prescription date, prescription days, and MEDD (Table 1). To calculate the overlapping MEDD for a certain intake date, a new table whose rows corresponded to each intake date was generated that transformed all the intake dates for a patient (i.e., not just the prescription dates) into rows by converting the prescription table (Table 2).

**Table 1.** Structure of a part of the narcotic prescription table.

| Row | Patient | Drug | Prescription date | Prescription day | MEDD |
| --- | --- | --- | --- | --- | --- |
| 1 | P1 | A | ****_**-01 | 5 | 15 |
| 2 | P1 | B | ****_**-01 | 7 | 30 |
| 3 | P1 | C | ****_**-02 | 5 | 20 |
| 4 | P2 | ... | ... | ... | ... |

Abbreviation: MEDD, morphine equivalent daily dose.

**Table 2.** Structure of a part of the modified narcotics prescription table according to each intake date to calculate overlapping MEDD.

| Row | Patient | Drug | Prescription date | Prescription day | MEDD | Intake date | Overlapping MEDD |
| --- | --- | --- | --- | --- | --- | --- | --- |
| 1 | P1 | A | ****_**-01 | 5 | 15 | ****_**-01 | 45 |
| 2 | P1 | A | ****_**-01 | 5 | 15 | ****_**-02 | 45 |
| 3 | P1 | A | ****_**-01 | 5 | 15 | ****_**-03 | 45 |
| 4 | P1 | A | ****_**-01 | 5 | 15 | ****_**-04 | 65 |
| 5 | P1 | A | ****_**-01 | 5 | 15 | ****_**-05 | 65 |
| 6 | P1 | B | ****_**-01 | 7 | 30 | ****_**-01 | 45 |
| 7 | P1 | B | ****_**-01 | 7 | 30 | ****_**-02 | 45 |
| 8 | P1 | B | ****_**-01 | 7 | 30 | ****_**-03 | 45 |
| 9 | P1 | B | ****_**-01 | 7 | 30 | ****_**-04 | 65 |
| 10 | P1 | B | ****_**-01 | 7 | 30 | ****_**-05 | 65 |
| 11 | P1 | B | ****_**-01 | 7 | 30 | ****_**-06 | 50 |
| 12 | P1 | B | ****_**-01 | 7 | 30 | ****_**-07 | 50 |
| 13 | P1 | C | ****_**-04 | 5 | 20 | ****_**-04 | 65 |
| 14 | P1 | C | ****_**-04 | 5 | 20 | ****_**-05 | 65 |
| 15 | P1 | C | ****_**-04 | 5 | 20 | ****_**-06 | 50 |
| 16 | P1 | C | ****_**-04 | 5 | 20 | ****_**-07 | 50 |
| 17 | P1 | C | ****_**-04 | 5 | 20 | ****_**-08 | 20 |
| 18 | P2 | ... | ... | ... | ... | ... | ... |

Abbreviation: MEDD, morphine equivalent daily dose.

A 3-month interval was set as the measurement interval for the time-series analysis (Jan-Mar, Apr-Jun, Jul-Sep, and Oct-Dec). The total number of prescriptions over 3 months was calculated. For the analysis of temporal changes in MEDD per patient, the highest MEDD during each 3-month period was selected and compared with the highest MEDD for the other 3-month periods.

R (R Foundation for Statistical Computing, Vienna, Austria. URL http://www.R-project.org/, ver. 3.6.0) and RStudio (RStudio, Inc., Boston, MA. URL http://www.rstudio.com/, ver. 1.2.1335) were used for statistical analyses. The ‘dplyr’ package in R was used to analyze data and the ‘ggplot2’ package was used to generate graphs. Outlier analysis was performed using the ‘outliers’ package. Cut-off value, sensitivity, specificity, and accuracy were calculated using the ‘pROC’ package. A two-tailed p value of less than 0.05 was considered statistically significant.

## RESULTS

### Patient and doctor outliers of the wt-MEDD score

This study involved 221 887 patients who were prescribed narcotics, 8737 doctors who wrote the prescriptions, and 555 097 narcotic prescriptions. After reviewing the 464 patients who were diagnosed with NUD in the CDM, only 29 patients were confirmed with NUD diagnosis following repeated narcotics prescription in our hospital. Most patients were diagnosed with NUD at other hospitals and transferred to our hospital (Fig. 2).

The cut-off wt-MEDD score for patient outliers (n=996) was 10.5 (*P* < 0.001). The list of doctor outliers (n = 23) for the wt-MEDD score is presented in Table 3 and Figure 3. This list of doctors and patients was extracted, and the hospital committee was notified to monitor prescription abnormalities and provide feedback to the relevant doctors.

**Table 3.** Partial list of doctor outliers of the wt-MEDD score.

| Rank | Doctor<br>(de-identified) | Department<br>(de-identified) | Prescription<br>number | Percentage<br>compared to<br>the mean<br>value (%) | Outlier<br>scores<br>(Z score) | Converted<br>P-value | Is outlier? |
| --- | --- | --- | --- | --- | --- | --- | --- |
| 1 | * | * | * | * | 60.678442 | < 0.000001 | TRUE |
| 2 | * | * | * | * | 39.274941 | < 0.000001 | TRUE |
| 3 | * | * | * | * | 33.336398 | < 0.000001 | TRUE |
| 4 | * | * | * | * | 26.614296 | < 0.000001 | TRUE |
| 5 | * | * | * | * | 13.994891 | < 0.000001 | TRUE |
| 6 | * | * | * | * | 12.469015 | < 0.000001 | TRUE |
| 7 | * | * | * | * | 8.056348 | < 0.000001 | TRUE |
| 8 | * | * | * | * | 6.612952 | < 0.000001 | TRUE |
| 9 | * | * | * | * | 5.788154 | < 0.000001 | TRUE |
| 10 | * | * | * | * | 5.540715 | < 0.000001 | TRUE |
| 11 | * | * | * | * | 5.375755 | < 0.000001 | TRUE |
| 12 | * | * | * | * | 5.252035 | < 0.000001 | TRUE |
| 13 | * | * | * | * | 4.757157 | 0.000002 | TRUE |
| 14 | * | * | * | * | 4.179798 | 0.000029 | TRUE |
| 14 | * | * | * | * | 4.179798 | 0.000029 | TRUE |
| 15 | * | * | * | * | 4.014839 | 0.000059 | TRUE |
| 16 | * | * | * | * | 3.973599 | 0.000071 | TRUE |
| 17 | * | * | * | * | 3.932359 | 0.000084 | TRUE |
| 18 | * | * | * | * | 3.808639 | 0.000140 | TRUE |
| 18 | * | * | * | * | 3.808639 | 0.000140 | TRUE |
| 19 | * | * | * | * | 3.767400 | 0.000165 | TRUE |
| 20 | * | * | * | * | 3.684920 | 0.000229 | TRUE |
| 21 | * | * | * | * | 3.643680 | 0.000269 | TRUE |
| 22 | * | * | * | * | 3.355001 | 0.000794 | TRUE |
| 23 | * | * | * | * | 3.313761 | 0.000921 | TRUE |

Abbreviation: wt-MEDD, weighted morphine equivalent daily dose.
\*Masked because it is sensitive information.

**Fig. 3.**
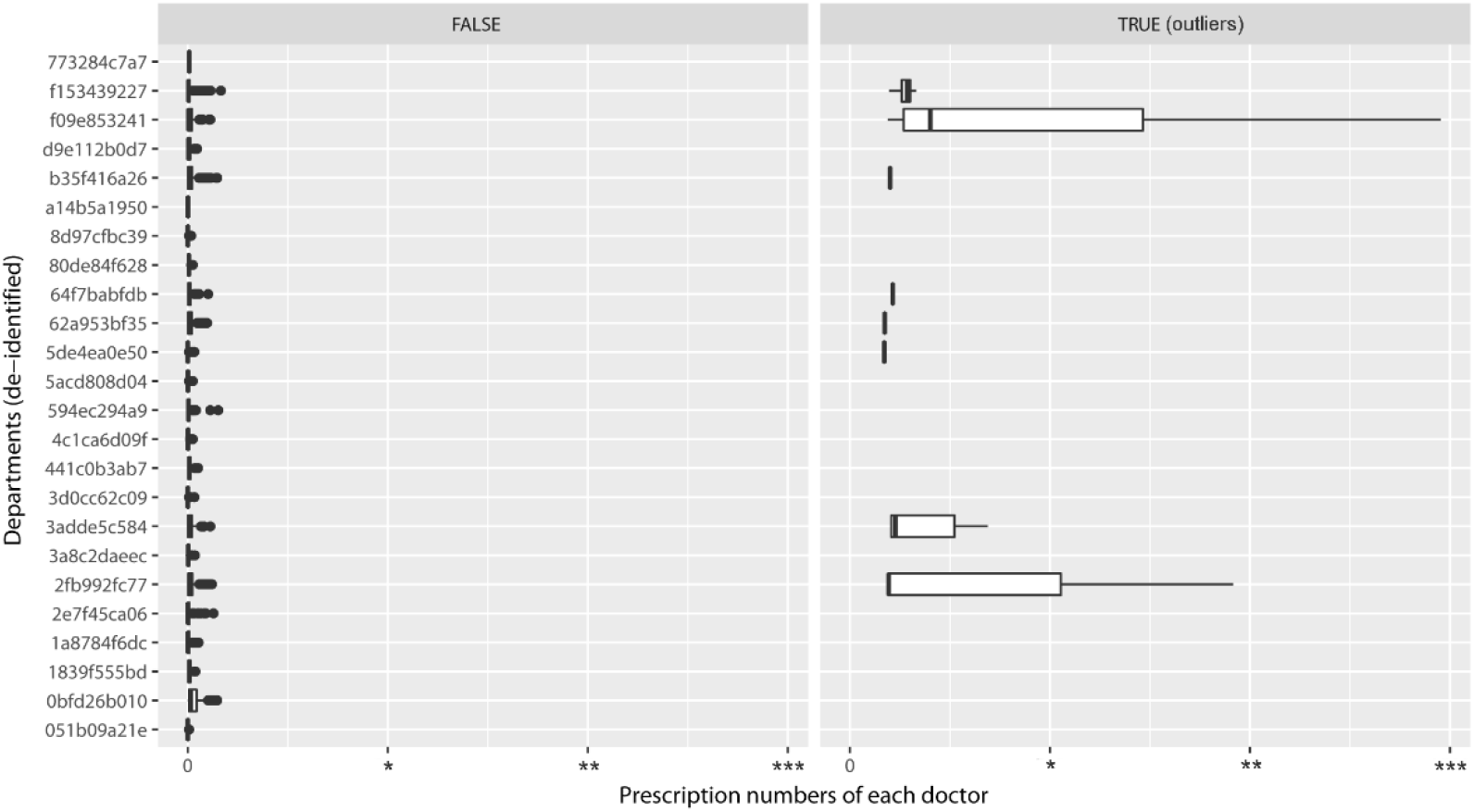
Outlier doctors for the weighted morphine equivalent daily dose score according to the departments.

### Comparison of wt-MEDD score and NUD high risk indexes to detect the diagnosed NUD

Table 4 compares the wt-MEDD scoring system and NUD high risk indexes for detecting diagnosed NUD. The optimal cut-off value for the wt-MEDD score with the highest sensitivity and specificity was ≥ 10.5, which was similar to the cut-off value for outlier wt-MEDD scores (*P* < 0.001). The median value of the wt-MEDD score among the 29 patients diagnosed with NUD was 52 (25–75th quantiles = 25–115), suggesting that most patients diagnosed with NUD had multiple prescriptions with a high MEDD ratio, higher than the prescription intention. Compared with other NUD high risk indexes, the wt-MEDD score showed the highest sensitivity and specificity (100.0% and 99.6%, respectively; Table 4). The McNemar test showed that the sensitivity and specificity of the wt-MEDD score were significantly superior to those of other indexes (*P* < 0.001), except for two indexes (quarter number with doctor number of ≥ 4 and number of prescriptions ≥ 10 days earlier than the scheduled visit) (Table 4).

**Table 4.** Comparison of the detection performance of various indexes for patients diagnosed with NUD.

| Indexes | Cut-off | Sensitivity (%) | Specificity (%) | Accuracy (%) | McNemar's P-value |
| --- | --- | --- | --- | --- | --- |
| **MEDD** |  |  |  |  |  |
| wt-MEDD score (*) | 10.5 | 100.0 | 99.6 | 99.6 | Reference |
| Highest overlapping MEDD (1) | 52.25 | 100.0 | 95.6 | 95.6 | <0.001 |
| Prescription number with intended MEDD $\geq 50$ | 10.5 | 93.1 | 99.3 | 99.3 | <0.001 |
| Prescription number with intended MEDD $\geq 90$ | 6.5 | 93.1 | 99.0 | 99.0 | <0.001 |
| **Frequency & duration** |  |  |  |  |  |
| Total prescription number (2) | 15.5 | 100.0 | 98.8 | 98.8 | <0.001 |
| Total quarter | 5.5 | 89.7 | 98.4 | 98.4 | <0.001 |
| Highest prescription number per quarter† for all prescription periods | 4.5 | 100.0 | 97.1 | 97.1 | <0.001 |
| **Prescription day** |  |  |  |  |  |
| Prescription number with $\geq 14$ prescription days at once | 11.5 | 89.7 | 99.5 | 99.5 | <0.001 |
| $\geq 30$ days at once (3) | 0.5 | 62.1 | 98.3 | 98.3 | <0.001 |
| $\geq 60$ days at once | 1.5 | 27.6 | 99.7 | 99.7 | <0.001 |
| $\geq 90$ days at once | 3.5 | 13.8 | 100.0 | 99.9 | <0.001 |
| **Number of doctors** |  |  |  |  |  |
| Total number of doctors (4) | 6.5 | 96.6 | 98.7 | 98.7 | <0.001 |
| Quarter number with doctor number of $\geq 4$ | 0.5 | 93.1 | 98.3 | 98.3 | 0.107 |
| Highest number of doctors per quarter† for all prescription periods | 2.5 | 96.6 | 96.3 | 96.3 | <0.001 |
| **Early prescription** |  |  |  |  |  |
| Prescription number with $\geq 1$ day earlier than scheduled visit | 1.5 | 93.1 | 99.1 | 99.1 | <0.001 |
| $\geq 2$ days earlier | 0.5 | 89.7 | 98.6 | 98.6 | <0.001 |
| $\geq 3$ days earlier | 0.5 | 89.7 | 98.8 | 98.8 | <0.001 |
| $\geq 4$ days earlier | 1.5 | 86.2 | 99.5 | 99.5 | <0.001 |
| $\geq 5$ days earlier | 1.5 | 86.2 | 99.5 | 99.5 | <0.001 |
| $\geq 6$ days earlier | 1.5 | 86.2 | 99.6 | 99.6 | <0.001 |
| $\geq 7$ days earlier (5) | 0.5 | 86.2 | 99.1 | 99.1 | <0.001 |
| $\geq 10$ days earlier | 1.5 | 82.8 | 99.7 | 99.7 | 0.548 |
| $\geq 14$ days earlier | 0.5 | 82.8 | 99.5 | 99.5 | <0.001 |
| **Combination of methods** |  |  |  |  |  |
| (1) ∩ (2) ∩ (4) (triple-test) | | 96.6 | 99.5 | 99.5 | <0.001 |
| (1) ∩ (2) ∩ (3) ∩ (4) | | 58.6 | 99.8 | 99.8 | <0.001 |
| (1) ∩ (2) ∩ (3) ∩ (5) | | 82.8 | 99.8 | 99.8 | <0.001 |
| (1) ∩ (2) ∩ (3) ∩ (4) ∩ (5) | | 58.6 | 99.9 | 99.9 | <0.001 |
| ((1) ∩ (2) ∩ (3) ∩ (4)) ∪ ((1) ∩ (2) ∩ (3) ∩ (5)) | | 82.8 | 99.8 | 99.8 | <0.001 |
| (1) ∩ (2) ∩ (4) ∩ (*) | | 96.6 | 99.7 | 99.7 | <0.001 |
| (1) ∩ (2) ∩ (3) ∩ (4) ∩ (*) | | 58.6 | 99.9 | 99.9 | <0.001 |
| (1) ∩ (2) ∩ (3) ∩ (5) ∩ (*) | | 82.8 | 99.8 | 99.8 | <0.001 |
| (1) ∩ (2) ∩ (3) ∩ (4) ∩ (5) ∩ (*) | | 58.6 | 99.9 | 99.9 | <0.001 |
| ((1) ∩ (2) ∩ (3) ∩ (4)) ∪ ((1) ∩ (2) ∩ (3) ∩ (5))) ∩ (*) | | 82.8 | 99.8 | 99.8 | <0.001 |

(\*) Abbreviations: MEDD, morphine equivalent daily dose.
†Quarters were divided as Jan-Mar, Apr-Jun, Jul-Sep, and Oct-Dec.

To improve the ability to detect NUD, we combined the NUD high risk indexes with the wt-MEDD score and calculated the sensitivity and specificity. A combined model using the highest overlapping MEDD, total prescription number, and total number of doctors (triple-test) showed excellent sensitivity and specificity (96.6% and 99.5%, respectively). When the wt-MEDD score and triple-test were combined, the sensitivity and specificity were 96.6% and 99.7%, respectively, suggesting that the wt-MEDD score can be used in combination with other NUD high risk indexes for screening patients with NUD (Table 4).

### Comparison of timepoints of reaching the wt-MEDD score cut-off value and that of the NUD diagnosis

In all 29 patients diagnosed with NUD, the timepoint when the wt-MEDD cut-off score was reached was earlier than the timepoint at which the doctor first diagnosed NUD. The mean duration of reaching the cut-off wt-MEDD score was 1024 days (median (Q1;Q3), 361 (192;2323)) from the initial narcotics prescription, while the mean duration until NUD diagnosis was 2578 days (median (Q1;Q3), 2342 (1396;4030)), resulting in a mean difference of 1554 days (95% confidence interval, 1096–2010 days, paired t-test, *P* < 0.001).

### Patient case report

To provide an example of the clinical application of this new methodology (wt-MEDD score) for the detection of NUD cases, one patient diagnosed with NUD who used multiple strategies to acquire a greater number of narcotics prescriptions was selected. Time-sequential changes in the wt-MEDD score and the NUD high risk indexes of this patient were retrospectively observed until the diagnosis of NUD (Fig. 4). Figure 4-(a) shows that the wt-MEDD score of the patient increased over time. The wt-MEDD score was 10 in November 2011, and the patient was diagnosed with NUD in October 2017. If the wt-MEDD score had been used as a screening method for this patient, he could have been diagnosed with NUD 6 years earlier.

**Fig. 4.**
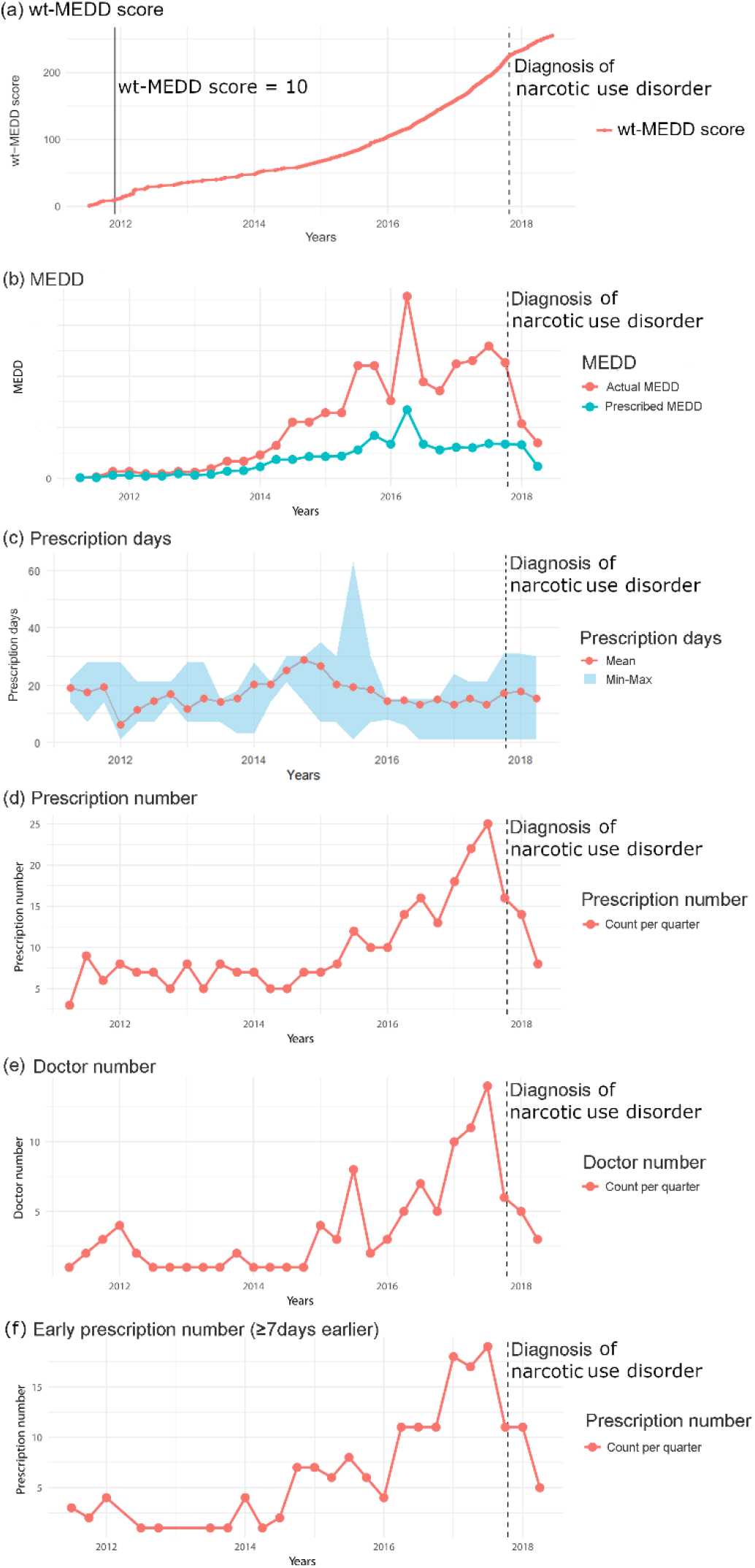
Temporal changes in various NUD high risk indexes for patients diagnosed with NUD. (a) wt-MEDD, (b) highest actual and intended MEDD every 3 months, (c) highest prescription days every 3 months, (d) total number of prescriptions every 3 months, (e) total number of prescribing doctors every 3 months, (f) number of early receipt of narcotics before ≥7 days for every 3 months. NUD, narcotic use disorders; MEDD, morphine equivalent; wt-MEDD, weighted MEDD.

The difference between the actual MEDD and intended MEDD (MEDD ratio) increased until the diagnosis of NUD (Fig. 4-b). The number of prescription days (mean, minimum, and maximum) increased just before the increase in the MEDD ratio (Fig. 4-c). The patterns of increase in the number of prescriptions (Fig. 4-d), number of prescribing doctors per 3-month period (Fig. 4-e), and early receipt of narcotics for more than 7 days (Fig. 4-f) were similar to the pattern of the increase in the MEDD ratio. This shows that use of multiple strategies can increase the MEDD ratio, and result in the diagnosis of NUD. Despite multiple strategies for receiving more narcotics, we could simply screen patients with NUD at an early prescription stage using only the wt-MEDD score without multiple indicators.

## DISCUSSION

The wt-MEDD score showed an impressive performance in detecting narcotic prescription patterns at an early stage in cases that were subsequently diagnosed with NUD. The wt-MEDD score corresponded to the number of prescription dates when the MEDD ratio was high. Patients with a wt-MEDD score of > 10.5 were identified as significant outliers, and this cut-off value was the same as the optimal cut-off value for detecting patients who had been diagnosed with NUD by doctors. The wt-MEDD score of 10.5 showed a high sensitivity (100%) and specificity (99.6%) in detecting patients diagnosed with NUD. These findings suggest that a wt-MEDD score of 10.5 could be used as a screening criterion to identify patients with NUD who have adopted various strategies such as “doctor shopping” for narcotics overuse.

Our hospital attempted to analyze abnormal patterns of narcotic prescription to prevent NUD in patients by providing feedback to the prescribing doctors. First, we referred to the CDC guideline ^7^, which is both reasonable and useful, as confirmed by the similarities with the optimal cut-off values used to detect patients with NUD in our study. However, the CDC guideline alone did not necessarily categorize a patient with NUD. Because we wanted to provide the prescribing doctors with information that more clearly reflected NUD, we considered situations that could be problematic with no exceptions. A patient possessing high doses of narcotics, which were not consistent with the intent of doctors’ prescriptions, could be regarded as a problematic situation and was therefore appropriate to be considered while defining NUD.

In the United States, the PDMP system automatically calculates the MEDD of prescription narcotics when narcotics are prescribed. The total MEDD of multiple prescriptions at the prescription date is provided; however, the overlapping MEDD owing to multiple prescriptions on a certain intake date is not calculated ^17^. Although the Ohio Automated Rx Reporting System reports daily MME on a screen for prescribers to view, this information is not stored and is therefore difficult to reconstruct ^17^. Several studies have analyzed overlapping prescriptions; however, they focused on overlapping benzodiazepine prescriptions with narcotics ^18^ or overlapping prescriptions alone rather than the overlapping MEDD ^19^. To the best of our knowledge, the present study is the first to have evaluated the wt-MEDD score, based on the number of prescription days when the MEDD ratio was high, as a tool for detecting abnormal prescription patterns.

The wt-MEDD score can be used to identify abnormal prescriptions, regardless of the strategy that the patient used to receive more narcotics. In addition, a graph showing the MEDD ratio over time can be provided for each patient in the outpatient clinic. The ability to immediately check for temporal changes by simply looking at the graph can dramatically shorten the time until the detection of unusual prescription patterns.

An opioid-risk tool, based on the fixed characteristics of patients, such as the history of alcoholism, has previously been reported ^20^. Although this opioid-risk tool has a primary prevention effect on NUD, it is not related to secondary prevention. In contrast, monitoring the wt-MEDD score enables screening before an increase in the number of abnormal prescriptions.

Our research findings have some limitations. Calculating the wt-MEDD score based on the overlapping MEDD could result in increased amounts of data, because every intake date (rather than only the prescription date) accounts for an additional row in the table. In our study, the data size of the table for each intake date was five times larger than that of the table for each prescription date. However, analyzing only patients who have been prescribed narcotics rather than all patients in the hospital would reduce the burden of data analysis. In addition, our study included only a small number of patients diagnosed with NUD, which might be attributed to the doctors being passive about diagnosing NUD. Particularly, for patients who have been prescribed narcotics in more than one department, it might be difficult to diagnose NUD owing to cross-department liability issues. Therefore, there may be more patients who could be diagnosed with NUD, especially among the wt-MEDD outlier group. Furthermore, we did not analyze narcotic prescriptions for patients with cancer, inpatients, and relatively low-dose narcotics (codeine and tramadol). These patients are expected to be analyzed using other criteria. In this study, we performed a retrospective analysis, and it is likely that a prospective trial will be needed to determine whether the number of patients with NUD can be reduced via monitoring of the wt-MEDD score. Finally, because our study population was from a single hospital, it did not consider the narcotics prescribed to patients at other hospitals. It is likely that patients with NUD will also be prescribed narcotics at other hospitals; therefore, it is necessary to introduce an integrated system that can check the records of narcotics prescriptions nationwide. In the case of the PDMP in the United States, all narcotic prescriptions for each patient are managed collectively at the state level ^21^. Furthermore, the diverse narcotic policies between hospitals and other countries might make such analysis difficult ^22^. However, if the system has the potential for detecting NUD via hospital prescriptions, a method for confirming the wt-MEDD score might be universally useful.

In conclusion, we defined the wt-MEDD score as the number of prescription days with a high MEDD ratio according to the definition of narcotic abuse. The wt-MEDD score detected patients diagnosed with NUD with a higher sensitivity and specificity than other metrics. Hence, monitoring the wt-MEDD score may facilitate early interventions for abnormal patterns of narcotics prescriptions by doctors and the development of NUD among patients.

## Data Availability

All data produced in the present study are available upon reasonable request to the authors

## ACKNOWLEDGMENTS

N/A

## AUTHOR CONTRIBUTIONS

YJK contributed to the conception, design of the work, acquisition, analysis, interpretation of data, and writing the manuscript. KHL contributed to the interpretation of data and substantively revised and wrote the manuscript. HRK, WSC, DYK, and YSK substantively revised the manuscript.

